# Real World Performance of the 21st Century Cures Act Population Level Application Programming Interface

**DOI:** 10.1101/2023.10.05.23296560

**Authors:** James R. Jones, Daniel Gottlieb, Andrew J. McMurry, Ashish Atreja, Pankaja M. Desai, Brian E. Dixon, Philip R.O. Payne, Anil J. Saldanha, Prabhu Shankar, Yauheni Solad, Adam B. Wilcox, Momeena S. Ali, Eugene Kang, Andrew M. Martin, Elizabeth Sprouse, David Taylor, Michael Terry, Vladimir Ignatov, the SMART Cumulus Network, Kenneth D. Mandl

## Abstract

**Objective:** To evaluate the real-world performance in delivering patient data on populations, of the SMART/HL7 Bulk FHIR Access API, required in Electronic Health Records (EHRs) under the 21st Century Cures Act Rule.

**Materials and Methods:** We used an open-source Bulk FHIR Testing Suite at five healthcare sites from April to September 2023, including four hospitals using EHRs certified for interoperability, and one Health Information Exchange (HIE) using a custom, standards-compliant API build. We measured export speeds, data sizes, and completeness across six types of FHIR resources.

**Results:** Among the certified platforms, Oracle Cerner led in speed, managing 5-16 million resources at over 8,000 resources/min. Three Epic sites exported a FHIR data subset, achieving 1-12 million resources at 1,555-2,500 resources/min. Notably, the HIE’s custom API outperformed, generating over 141 million resources at 12,000 resources/min.

**Discussion:** The HIE’s custom API showcased superior performance, endorsing the effectiveness of SMART/HL7 Bulk FHIR in enabling large-scale data exchange while underlining the need for optimization in existing EHR platforms. Agility and scalability are essential for diverse health, research, and public health use cases.

**Conclusion:** To fully realize the interoperability goals of the 21st Century Cures Act, addressing the performance limitations of Bulk FHIR API is critical. It would be beneficial to include performance metrics in both certification and reporting processes.

## INTRODUCTION

The SMART/HL7 FHIR Bulk Data Access API [1] is designed to enable standardized access to electronic health records (EHRs) on large populations of patients. The API facilitates the “push button population health” thus fostering data-driven innovation on local, regional, and national scales. As of December 31, 2022, support for it is required universally. The 21st Century Cures Act [2] stipulates that certified health information technology must incorporate an application programming interface (API) that provides access to all data elements of a patient’s electronic health record, in a manner requiring no “special effort.” In the spring of 2020, a regulation—the 21st Century Cures Act Interoperability, Information Blocking, and the ONC Health IT Certification Program [3], were introduced by the Office of the National Coordinator of Health Information Technology (ONC) to govern the API prerequisite, while also ensuring safeguards against information blocking. This rule requires support for the SMART/HL7 FHIR Bulk Data Access API to allow access to patient-level data across a population, bolstering an array of applications across healthcare, research, and public health. Data exchanged are over 100 elements defined by the US Core Data for Interoperability (USCDI) [4] in Fast Health Interoperability Resources (FHIR) format.

To comply with the Centers for Medicare and Medicaid Services (CMS) Promoting Interoperability Programs [5] and avoid a negative payment adjustment, use certified health information technology (IT). Developers of IT get certified by an authorized certification body that tests for technical compliance but does not currently measure API performance. Specifically, the 170.315 (g) (10) certification criterion is a federal mandate stipulating that health IT developers must offer standardized APIs for individual services [6] and for population-based services [7]. This criterion is included in the 2015 Edition Base EHR definition, as part of the Cures Act Final Rule [4] .

The Bulk FHIR Access API is new in 2023 and this exploratory work sought to test these early implementations to understand current state and then possibilities as implementations are iterated on over time. Any newly deployed software requires real world testing to benchmark performance. We measured Bulk FHIR Access API performance across a collaborative network of five provider sites using a range of technologies, spanning two EHR vendor products and a health information exchange (HIE) data repository.

## METHODS

### Participants

We selected five healthcare sites with high performing information technology teams from across the US. All were university-affiliated academic medical centers. The FHIR APIs at three sites were provided by Epic (one was remote hosted) and one by Cerner (also remote hosted) as part of their g(10) certified products. The HIE fashioned its own Bulk FHIR Access API as a facade on top of a local relational database populated by HL7 V2 messages. Following the Bulk FHIR Access Implementation Guide, the HIE service reads rows from its relational database in bulk, converts them into FHIR resources, writes them into NDJSON files, and returns URLs for downloading them to a final Bulk FHIR polling request upon completion.

### Bulk FHIR Testing Suite

We relied on the open source SMART Bulk FHIR Client [1,8], which each performance site downloaded and set up behind their institutional firewall, creating a local implementation. As part of this effort, the Bulk Data Client was instrumented to log the elapsed time for each component of the data retrieval and download workflow.

### FHIR resource types in scope

The FHIR data format employs a modular approach, breaking down healthcare information into an array of data models called “Resources” that range from medical conditions to medication requests, each with its distinct searchable parameters and governance rules. As a result, every patient’s healthcare journey is represented by a unique combination of these Resource types, which can be selectively accessed as needed. We focus on six specific Resource models—Patient, Encounter, Condition, DocumentReference, Observation, and MedicationRequest—that align with the criteria set forth in the USCDI v1 standard and are pertinent to population health scenarios.

### Cohort creation

The Cures Act Final Rule [3] requires certified EHRs to support exporting pre-defined groups of patients through the API described in the Bulk FHIR specification. Notably, the process with which healthcare sites define these groupings of patients within the EHR and the supported group size are not standardized across systems. Cerner currently places a limitation of 20,000 patients per list and suggests groups are kept at or below 10,000 patients for performance reasons. For group attribution in Cerner, the process begins with a query using Cerner’s Discern Visual Developer tool. Next, the resulting Cerner PERSON_IDs are transcribed to a spreadsheet, which is uploaded into the Ignite Management Tool. For the duration of this project, the Ignite Management Tool was unavailable; however, the site was able to work with Cerner for assistance.

Epic uses pre-existing registry functionality that does not place a limitation on the number of patient records that can belong to a group but encourages clients to limit requests to groups at or below 1,000 patients. Both Epic and Cerner provide the ability to define patient groups by inclusion criteria such as presence of specific coded diagnoses or having a visit of a specific type within a time window, though the interfaces differ. To compare performance across varied EHR vendor implementations and varied hospital volumes, a set of adaptable inclusion criteria was selected to define patient groups of varying sizes between 1,000 and 20,000. The HIE has a dedicated Data Core team that provisioned their testing team with a single large list of patients based on a current public health project leveraging Bulk FHIR. A breakdown of the sites and patient groups involved in each test is provided in **Table 1**.

**Table 1.** Description of sites contributing performance data to the study.

| Site (API developer) | Server location | g(10) certified | Bulk API filtering options | Records / max group size | Group selection criteria and notes |
| --- | --- | --- | --- | --- | --- |
| Site 1 (Cerner) | Remote Hosted | Yes | _type<br>_typeFilter<br>_since | <10k / 20k | • Patients with an emergency department visit during the month of January 2023<br>• Patients with an emergency department visit during Q3 2021. Only pediatric population in the test.<br>• Patients with an emergency department visit during Q4 2022. Only pediatric population in the test. |
| Site 2 (Epic) | On Premises | Yes | _type | <1k / No limit | • Patients with an emergency department visit during the most recent week<br>• Patients with an emergency department visit during the most recent month |
| Site 3 (Epic) | Remote Hosted | Yes | _type | <1k / No limit | • Patients with an emergency department visit during the most recent week<br>• Patients with an emergency department visit during the most recent month |
| Site 4 (Epic) | On Premises | Yes | _type | <1k / No limit | • Hypertension management Tier 5 cohort<br>• Asthma Utilization Tier 4 cohort<br>• Congestive Heart Failure cohort |
| Site 5 (HIE Custom) | On Premises | No | _type<br>_since | No limit | • Patients with a positive COVID diagnosis in the county. Exports were done 1 month at a time using “_since” and an experimental “_until” parameter. |

### FHIR API Queries

Each site sought to extract at least one year of data on the populations slated for testing, across six FHIR resource types. Sites were encouraged to limit the observation resource categories to only those for laboratory results, vital signs measurements, and smoking status records. Multiple API parameters were defined in the bulk data access implementation guide, and sites were encouraged to use the “_type” parameter to limit resource type requests.

Date filtering could be accomplished either by the “_since” bulk API parameter for timestamps or the more flexible “_typeFilter” parameter that enables clients to provide FHIR REST search queries. Uniform date filtering was not required to accommodate variations in Bulk FHIR implementations. The testing plan was circulated in January 2023 to the five participating sites and teams managing access to their Bulk FHIR APIs, with a goal to review results in March.

### SMART on FHIR Testing Suite

As an alternative approach to extracting a cohort from the EHR FHIR Bulk Data API, we tested the submission of many repeated requests using the SMART on FHIR API with a “Crawler” client developed for the task that iterated over the given population one patient at a time. Of note, the single patient FHIR API has a different set of configurable search parameters, allowing sites that were not able to use the “_since” or “_typeFilter” parameters to request a more targeted population for comparison.

## RESULTS

All sites were able to install local copies of the Bulk FHIR Testing Suite and four of the five sites were able to perform a series of tests between April and August of 2023. The four sites using g(10) certified EHR products each used the vendor provided tools to request groups be provisioned for the purpose of performance testing. Sites encountered some issues and at times substantial delays as they learned to navigate these tools. It took between two and 119 days (on average, 65 days) for sites to successfully make any Bulk FHIR API requests after providing selection criteria. The sites required coordination with one or more teams outside of their departments or organizations to gain sufficient access to run the test suites. Notably, this was, for most sites, the first project provisioning Bulk FHIR APIs, and new configurations and workflows involving multiple parties needed to be established. Errors were encountered in the tooling provided by the certified technology, which took time to report to the EHR developers and for them to identify and implement solutions. A breakdown of all the test exports is in **Table 2**.

**Table 2.** Benchmark results. Number of resources exported in each test, with measurements of total time, resources per minute, and seconds needed to export one patient’s complete record on average. P = Patients, E = Encounters, O = Observations, C = Conditions, D = Document References, M = MedicationRequests, R = total Resources. SoF = SMART on FHIR testing suite.

| Site | Client | P | E | O | C | D | M | R | Total hours | Resources per min. |
| --- | --- | --- | --- | --- | --- | --- | --- | --- | --- | --- |
| Site 1 (Cerner) | SoF | 4,376 | 180,971 | 4,365,361 | 97,117 | Errors - 0 | 347,605 | 4,995,430 | 13.1 | 6349.9 |
| Site 1 (Cerner) | Bulk FHIR | 4,376 | 180,971 | 4,366,797 | 97,116 | 301,078 | 347,593 | 5,297,931 | 8.1 | 10,838.3 |
| Site 1 (Cerner) | Bulk FHIR | 10,244 | 541,226 | 11,701,214 | 271,617 | 1,494,026 | 973,404 | 14,991,731 | 34.2 | 7,300 |
| Site 1 (Cerner) | Bulk FHIR | 13,462 | 547,811 | 12,577,800 | 304,693 | 1,659,861 | 991,942 | 16,095,569 | 32.5 | 8,261 |
| Site 2 (Epic) | SoF | 892 | 169,902 | 1,058,451 | 256,113 | 214,654 | 136,098 | 1,836,110 | 5.9 | 5,186.8 |
| Site 2 (Epic) | Bulk FHIR | 907 | 181,493 | 1,448,415 | 173,754 | 461,601 | 150,076 | 2,416,246 | 25.9 | 1,554.9 |
| Site 2 (Epic) | SoF | 2,686 | 1,104,900 | 6,103,568 | 1,502,393 | 1,286,443 | 726,654 | 10,726,644 | 73.4 | 4,792.2 |
| Site 3 (Epic) | SoF | 1,173 | 250,879 | 988,092 | 333,917 | 327,839 | 365,195 | 2,267,095 | 4.6 | 8,214.1 |
| Site 3 (Epic) | Bulk FHIR | 1,269 | Errors - 0 | 3,978,688 | 347,782 | Errors - 0 | 399,573 | 4,727,312 | 27.9 | 2,826.8 |
| Site 3 (Epic) | Bulk FHIR | 4,217 | Errors - 0 | Errors - 0 | 1,165,583 | Errors - 0 | 1,465,153 | 2,634,953 | 20.3 | 2,163.3 |
| Site 4 (Epic) | SoF | 1,021 | 16,737 | 61,633 | 209,533 | 20,723 | 12,117 | 321,764 | 6.8 | 788.6 |
| Site 4 (Epic) | Bulk FHIR | 1,020 | 153,684 | 661,101 | 134,102 | Errors - 0 | 88,870 | 1,038,777 | 34.5 | 501.8 |
| Site 4 (Epic) | Bulk FHIR | 5,059 | 2,064,125 | 7,611,121 | 1,915,051 | Errors - 0 | 1,212,564 | 12,807,920 | 83.7 | 2,550.4 |
| Site 4 (Epic) | SoF | 8,311 | 1,261,097 | 4,374,770 | 4,363,954 | Errors - 0 | 1,803,446 | 11,811,578 | 90.2 | 2,183.2 |
| Site 4 (Epic) | Bulk FHIR | 10,189 | 3,863,233 | 33,667,978 | 3,867,079 | Errors - 0 | 3,961,808 | 45,370,287 | 330 | 2,291.4 |
| Site 5 (HIE custom) | Bulk FHIR | 2,403,820 | 11,497,279 | 92,479,812 | 11,902,665 | 23,225,023 | 94,308 | 141,602,907 | 215.8 | 12,215.7 |

### Time taken to export records by server and method

Logs created by the Bulk FHIR Testing Suite and the SMART on FHIR Testing Suite were analyzed to generate statistics comparing the performance of the local implementations. Statistics include total counts of each resource type able to be downloaded for the cohort, total time taken, and overall number of resources able to be exported and downloaded per minute.

### Performance scaling differences between vendors

In this initial series of tests on some of the earliest Bulk FHIR services in production, the Oracle Cerner implementation demonstrated higher overall Bulk FHIR export speeds than the three Epic implementations, and both were slower than the HIE bulk service. Of course, these test sites represent only a small sample of certified implementations from both vendors, and do not permit control for differences among EHR implementations. Tests were run on populations larger than Epic’s recommended threshold of 1,000 patients per request. As seen in **Figure 1**, the custom-built HIE FHIR Service performed very well, averaging over 11,000 resources per minute over its export, and the Oracle Cerner site averaged over 8,000. On these three early Epic Bulk FHIR implementations, export speeds were highly variable on the smaller tests and were observed to approach 2,500 resources per minute on groups larger than 1,000 patients. Discussion with Epic engineers on the performance for larger groups inspired our development and use of the SMART on FHIR test suite that allowed sites to make serial requests “one patient at a time”. This method was intended to showcase a baseline of performance and to allow additional filtering options initially unsupported by Epic’s Bulk APIs. Interestingly, early tests showed instances of this approach providing Epic sites faster and more complete access to bulk exports of interest than they could obtain through their Bulk FHIR APIs. In contrast, the Cerner implementation slowed down when resources were requested one patient at a time with this method.

**Figure 1.**
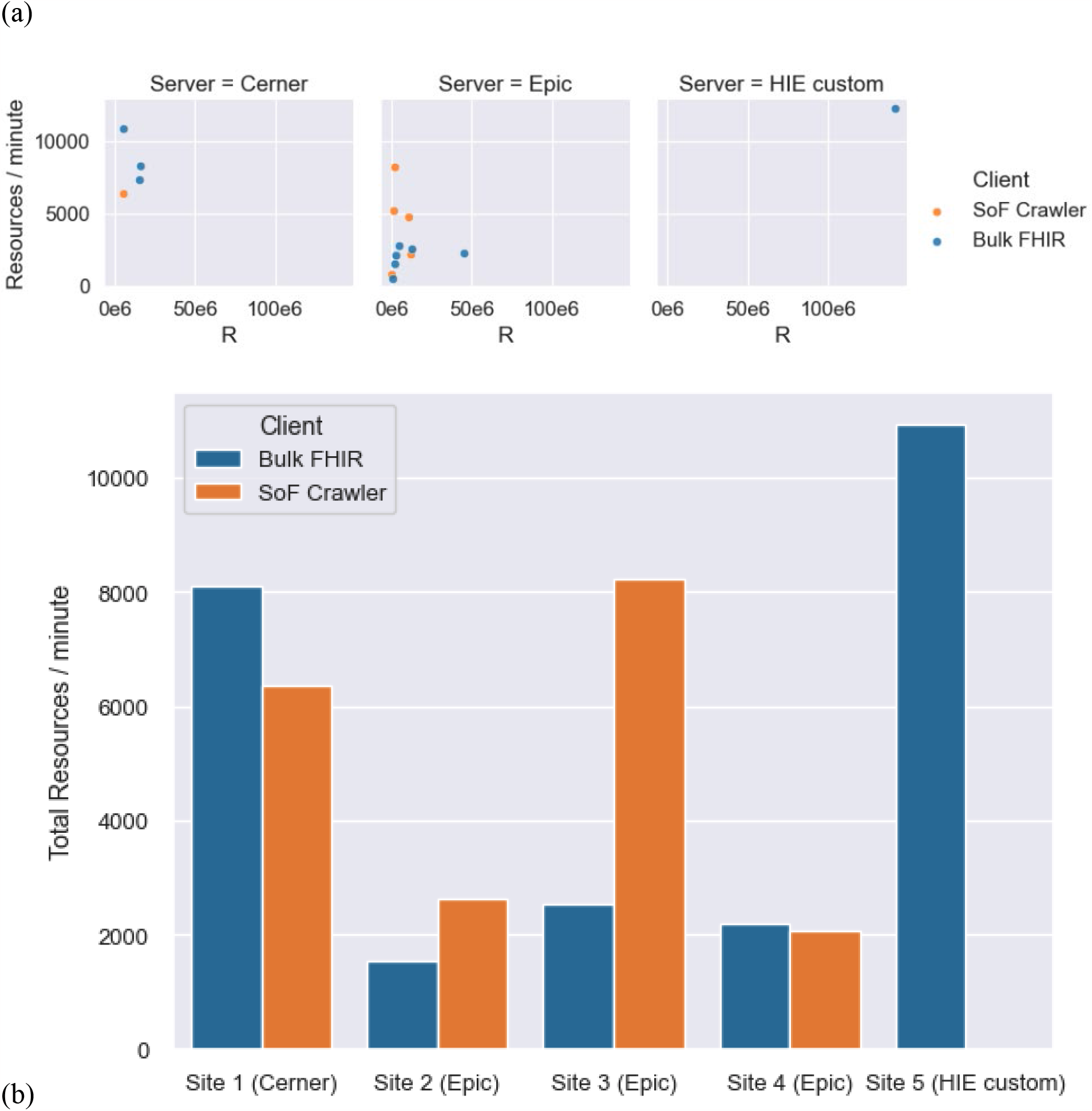
Performance scaling differences between vendors. **(a)** Measured export speed plotted against total number of resources R in each benchmark. **(b)** Total count of resources exported at each site divided by total export time. Performance is grouped by API service provider.

### Performance scaling differences between implementations from a single vendor

Sites 2, 3, and 4 each used APIs provided by Epic, and showed a range of performance in the tests, between 1555 and 2500 resources per minute. The throughput and number of resources that were able to be exported for individual requests for these sites are shown in **Figure 2**. Site 3 split the tests up into requests for individual resource types and was able to complete some of the exports on a cohort of over 4,000 patient records. Site 4 limited their requests to not include Document References and was then able to successfully export for groups of 1,000, 5,000 and 10,000 patient records.

**Figure 2.**
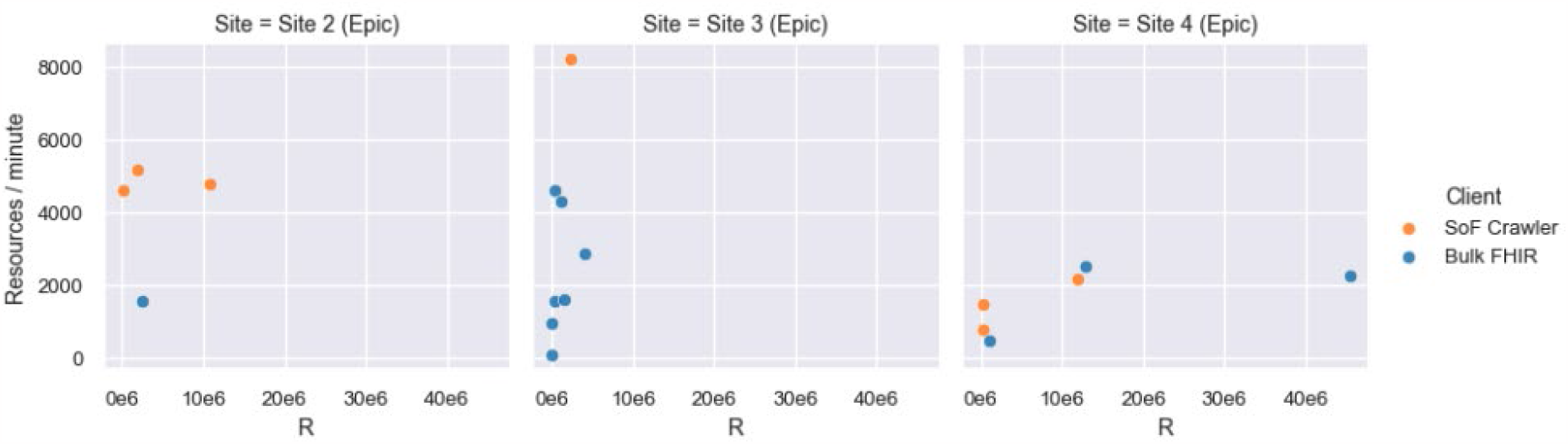
Performance scaling differences between implementations from a single vendor.

### Resource Volume and Performance by Resource type

Large volume and performance variances were measured across resource types. Among the six FHIR Resource models in this test, Observation stands out as both the fastest and most abundant Resource for a majority of patients. It offers a flexible data structure that encompasses specific metrics such as laboratory results and vital statistics. As can be seen in **Figure 3**, averaged over all the servers and tests, patients had around 2,000 linked observations each when exported without further filtering, compared to around 300 each of the other resource types in the study. Observation is also on average one of the smallest resource types, averaging 2KB per instance. Only the Condition resource that carries coded diagnosis information was measured to be smaller in average size. Of note, the DocumentReference resource carries metadata about documents including clinical notes, often present only in the resource through a reference to another “Binary” Resource. The Binary endpoint containing the clinical note text was not directly tested.

**Figure 3.**
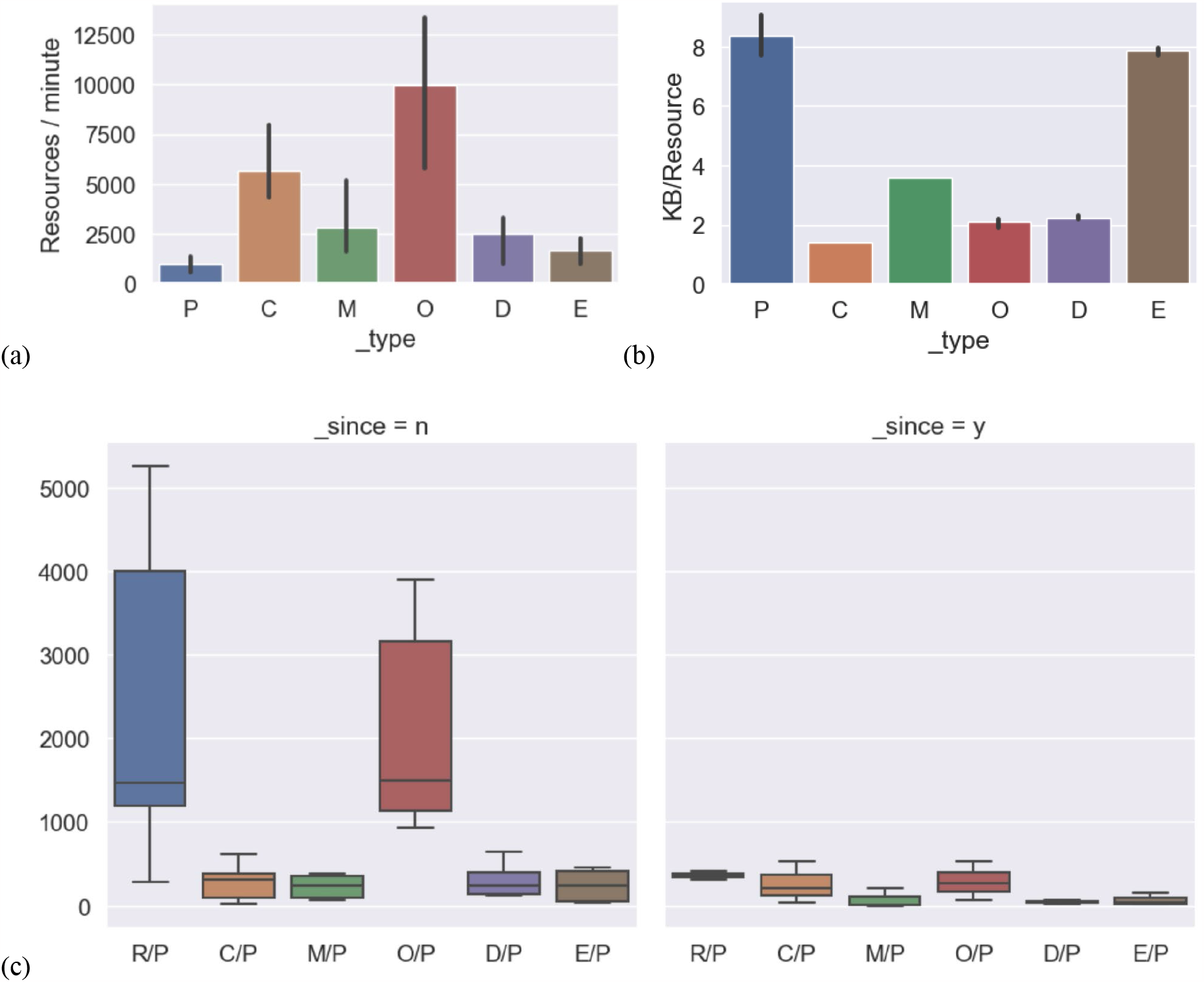
Effects of bulk API parameters. (a) average export speeds by resource type, seen by altering the “_type” parameter. (b) breakdown of average resource size for each type in the study, in kilobytes. (c) average number of resources of each type per patient in the query. Counts on the left are without use of the “_since” parameter, and counts on the right are from queries using the “_since” parameter to only export data from the last year or as simulated by date filtering in the SMART on FHIR testing suite. P=Patients, E=Encounters, O=Observations, C=Conditions, D=Document References, M=MedicationRequests, R=total Resources.

Other filters were employed where supported to narrow the request to only resources of interest. Using “_since”, “_typeFilter”, or corresponding date filters from the SMART on FHIR test suite enabled requesting only 1 year of data, which significantly reduced the number of resources in the exports as shown in Figure 3.c.

### Challenges

The endpoints at Sites 1, 2, and 5 were able to complete exports that included the six resource types using the Bulk FHIR test suite. Sites 3 and 4 (both Epic) encountered errors that required fallback to the SMART on FHIR REST API to export some of the resource types successfully:Document References and Encounters at Site 3 and Document Referencesat site 4.

Some errors were encountered that were determined to be caused by data incompleteness or improper template configuration. For example, at Site 1 (Cerner), data elements required by the FHIR API were missing or not represented to the vendor as expected. This prevented the site from reliably exporting all Observation and DocumentReference instances for the cohorts over the study period without falling back to the patient by patient SMART on FHIR REST API to fill in gaps. Over two months, Cerner worked with the site to troubleshoot the issue and provide better error handling, which enabled exports to finish that contained errors, rather than having them fail prematurely and return none of the previously generated data. The limitation that patient groups are capped at 20,000 introduces additional workflow complexity to manage exports over a larger population. Lastly, while Cerner does support date filtering with the “_since” bulk API parameter, early tests showed no decrease in the amount of time the server took to prepare and complete filtered vs unfiltered requests.

Among the initial Epic implementations tested, successfully exporting Bulk FHIR for groups of 1,000 patients or more was difficult, with additional issues being identified as requests scaled. Several exports ran into errors and had to be aborted after multiple days of processing. After careful configuring, sites 3 and 4 were able to export a subset of resource types for groups of around 5,000 patients. Site 4 worked closely with Epic to investigate issues and discuss the use cases being tested and possible enhancements. They were then able to complete an export request for over 10,000 patients, though the export took nearly two weeks to complete and an expired token prevented downloading the data in the end.

## DISCUSSION

In the realm of 21st Century interoperability regulatory science, these measurements represent the first performance evaluations, to our knowledge, of early SMART/HL7 Bulk FHIR API implementations. This study served a dual purpose: it not only provided critical performance metrics but also catalyzed robust discussions and led to tangible improvements in software and workflows among healthcare systems, EHR vendors, and API end users.

Because the custom API implementation at the HIE, which took only a few weeks to design, implement and use, outperformed the g(10) certified implementations, substantial potential for improvement by EHR vendors is evident. Our experience highlights that for this technology to reach its full potential, and indeed meaningful compliance with the law and regulation, more work is needed on the EHR vendors’ Bulk FHIR implementations. ONC sponsored two meetings, one before the Rule’s publication and one after [9,10], to identify high value use cases for Bulk FHIR. While the vendors have now stood up and supported initial implementations, real world performance of the APIs is not quite there yet to support many of these use cases. The barriers persist even when EHR vendor support is available to high-performing IT teams at leading care delivery sites.

To address myriad identified use cases in care delivery, value, research and population health, meaningful compliance with the Cures Act will require EHR developers to sustain focus on improving current implementations. In some cases, we found that making thousands of individual FHIR requests using the SMART crawler was faster than making a single request for the same data in bulk, which indicates there may be sequential processes in the bulk API response that could be parallelized. Cerner supports the inclusion of parameters in bulk requests to filter and limit the FHIR data being returned. Epic does not yet support these filters, which could improve performance by bypassing the processing of unnecessary results. The highly performant bulk implementation at the HIE, built on their data warehouse, suggests that all EHR vendors could potentially achieve substantial improvements, as Cerner does now, by investing in bulk export capabilities that run on their data warehouses rather than their transactional databases.

With the current performance limitations, large enough datasets for some use cases could take not hours but years to be generated. Although Epic has suggested limiting the use of the Bulk FHIR Access API to 1,000 patients or fewer, we believe that such restrictions are inconsistent with the intent of the API provisions of the Cures Act Rule. Incorporating performance measurements alongside technical compliance to incentivize and track real world usability of the APIs should be considered both in the ONC Health IT Certification Program [11] and the EHR reporting program [12]. Yet, interesting challenges lie ahead in designing a performance-based criterion. For instance, being able to export a flat number of records per day won’t scale for larger health systems that may have much more patient volume. A specific metric may be needed, for example the ability for health systems to export US Core data for their entire active patient population within a certain time frame, say 5 days. All interested parties—the EHR vendors, healthcare organizations, policymakers—can work together to establish feasible metrics. Partnering in this discussion will allow everyone to collaborate on this imperative technology.

This study is intended as a contribution to the regulatory science around health IT, informing the development and refinement of interoperability law and regulation. Future investigations could broaden the scope by including additional FHIR resource types, engaging more sites, and incorporating a wider array of vendors. To sharpen the performance assessment, subsequent tests might examine server response times for various request stages, and compare peak and off-peak speed variations. The standout performance at the HIE site underscores a compelling avenue for innovation: the development of population-level Bulk FHIR interfaces. These could synergize with patient data soon to be accessible using the Trusted Exchange Framework and Common Agreement (TEFCA), thereby unlocking new dimensions of data usability and access [13].

To achieve the seamless healthcare data exchange envisioned by the 21st Century Cures Act, where all elements of a patient’s record can be accessed across an API with ‘no special effort,’ a unified and urgent effort from healthcare providers, EHR vendors, and policymakers is crucial. While testing came with its challenges, the learnings from this effort will allow continued, strong partnerships with certified health information technology vendors as key interested parties work to transform clinical and public health informatics. The experience of our collaborative network reinforces the value the Bulk FHIR API has as a transformative tool.

## Data Availability

All data produced in the present study are available upon reasonable request to the authors.

## ACKNOWLEDGEMENTS

This study was supported by the Office of the National Coordinator of Health Information Technology (contract 90AX0031); the Centers for Disease Control and Prevention of the U.S. Department of Health and Human Services (HHS) as part of a financial assistance award (The contents are those of the author(s) and do not necessarily represent the official views of, nor an endorsement, by CDC/HHS, or the U.S. Government); the National Center for Advancing Translational Sciences (cooperative agreement U01TR002623); the National Association of Chronic Disease Directors/Centers for Disease Control and Prevention (grant NU38OT000286); and the Centers for Disease Control and Prevention (grant U18DP006500, cooperative agreement NU58IP000004). We acknowledge the participation of Regenstrief Institute, Inc. in this project. The content is solely the responsibility of the authors and does not necessarily represent the official views of the funding sources.

## Notes

### Competing Interest Statement

The authors have declared no competing interest.

### Author Declarations

The Institutional Review Board of Boston Children's Hospital waived ethical approval for this work.

### Summary of Updates

Minor corrections.

